# Prevalence of diabetes among Indigenous women in Guatemala: A retrospective chart review study of 13 643 patients between 2015-2022

**DOI:** 10.1101/2024.01.08.24300932

**Authors:** Stephen Alajajian, Jenny Bartolimin, Yolanda Juarez Martin, Caitlin Scott, Peter Rohloff, David Flood

**Author notes:** Corresponding author Stephen Alajajian, Present address: 1923 Jefferson St. Baltimore, MD 21205.

## Abstract

There are limited data on diabetes among Indigenous populations in Guatemala. In a retrospective chart review of a clinical program serving more than 13 000 primarily Indigenous women in Guatemala, age-adjusted diabetes prevalence was 7.9% (95% CI: 7.3 to 8.5), and 37.9% (95% CI: 35.1 to 40.8%) of women were undiagnosed.

## INTRODUCTION

Guatemala is a middle-income, Central America nation in which nearly half of the population are Indigenous.^1^ Modeling studies suggest that the diabetes burden is rising rapidly in Guatemala.^2,3^ However, like other Latin American countries with large numbers of Indigenous people, in Guatemala there is limited data on the epidemiology of diabetes in this marginalized population. The objective of this study is to investigate the prevalence of diabetes in a clinical sample of primarily Indigenous women in Guatemala.

## METHODS

We conducted a retrospective chart review using data from a non-governmental, community-based primary care program serving adult women in nine Central and Western departments in Guatemala. Program details previously have been described.^4,5^ Upon enrollment in the program, patients were queried if they had a history of diabetes and, if not, were screened for diabetes using random or fasting glucose capillary measurements. For this analysis, we extracted electronic health record and administrative data between July 1, 2015, and December 31, 2022. We defined diabetes by (1) self-reported prior history, (2) current use of a glucose-lowering medication, or (3) an elevated blood glucose (fasting ≥126 mg/dl or random ≥200 md/dl) using the last available recorded glucose value.

We calculated diabetes prevalence in the overall sample and by patient characteristics including body mass index (BMI) categories, ethnicity, language, residence, and economic status. We age-standardized estimates to the World Health Organization (WHO) reference population.^6^ Economic status was assessed using the Poverty Probability Index, which uses asset ownership to predict the probability that a household was below the national poverty line.^7^ Absolute differences and relative differences (as risk ratios) in diabetes prevalence were computed within levels of each characteristic. We used multiple imputation with chained equations to impute missing data on patient characteristics (50 iterations). Results were reported with logit-transformed 95% confidence intervals.^8^ We used Stata version 17. Ethics approval was obtained from the Wuqu’ Kawoq Institutional Review Board (WK-2022-004).

## RESULTS

The final analytic sample was 13,643 patients after excluding those with no glucose data recorded (n=387) and without a legitimate medical record number (n=3). Among the sample, the last available glucose was fasting glucose for 2,172 women (16.0%) and was random glucose for 11,471 women (84.1%). The median age was 36 years (interquartile range [IQR]: 28-47). The median BMI was 27.3 kg/m^2^ (IQR: 24.1-30.9); 38.7% of the sample were overweight, and 30.2% were obese. The sample was predominantly Indigenous (90.3%) and lived in rural areas (85.1%). Overall, 44.4% of the sample were living in poverty. Missingness was high for area of residence (43.9%) and economic status (42.3%).

Across the overall sample, there were 1,126 women with diabetes. The crude diabetes prevalence was 8.3% (95% Confidence Interval [CI]: 7.8 to 8.7), and the age-adjusted diabetes prevalence was 7.9% (95% CI: 7.3 to 8.5). Among those with diabetes, 62.1% (95% CI: 59.2 to 64.5%) of women self-reported a prior diagnosis and 37.9% (95% CI: 35.1 to 40.8%) of women were newly diagnosed.

By patient characteristics, diabetes prevalence rose sharply with increasing age, reaching nearly 20% in women who were 50 years of age or older. Compared to the reference of normal BMI, women who were obese had an age-standardized diabetes prevalence that was 3.2% (95% CI: 1.1 to 5.2) greater in absolute magnitude and approximately 40% greater in relative magnitude (risk ratio: 1.4 [95% CI: 1.1 to 1.8). Age-standardized diabetes was higher among non-Indigenous versus Indigenous women and among Spanish versus Mayan language speakers. Compared to those most likely to be in poverty, women least likely to be in poverty had an age-standardized diabetes prevalence that was 6.0% (95% CI: 3.7 to 8.3) greater in absolute magnitude and 100% greater in relative magnitude (risk ratio: 2.0 [95% CI: 1.5 to 2.6).

## DISCUSSION

In a retrospective chart review of a clinical program serving more than 13 000 primarily Indigenous women in Guatemala, we found that the diabetes prevalence was approximately 8% and that approximately 38% of people with diabetes were undiagnosed. Across patient characteristics, women who were obese had a 40% higher risk and women in the highest economic quartile had nearly a 2-fold higher risk of having diabetes. At the same time, diabetes prevalence was sizeable even among women who were Indigenous (8.4%), lived in rural areas (8.1%), or were in the lowest economic quartile (5.9%).

To our knowledge, this study leverages the largest programmatic or survey dataset of primarily Indigenous people with diabetes in Guatemala. A few prior household surveys conducted in select predominantly Indigenous areas have reported diabetes prevalence ranging from 3.0% to 13.5%.^9-11^ Differences in these survey estimates may be explained by variations in diabetes definition, time period of assessment, or differences in the underlying population. A national survey conducted from 2018 to 2019 among approximately 2,500 reproductive aged women aged 15-49 years reported diabetes prevalence of approximately 6%.^12^ Unfortunately, to date, there is no other Guatemalan national health survey—such as a WHO STEPS survey—that includes diabetes assessments.

The strength of the current study is our large sample size and use of repeat testing to confirm diabetes among newly diagnosed women with elevated random glucose. The large number of women in our sample gives us added power to investigate differences in diabetes prevalence across meaningful subgroups of the population. The main limitation of the study is that our sample of patients is restricted to women and was not drawn from a representative sampling frame. At the same time, our clinical sample is quite similar in population age structure, language preference, and economic status as the overall Indigenous population of women in these geographic areas of Guatemala.^13^

In conclusion, we report diabetes prevalence of diabetes approximately 8% in a large clinical sample of Indigenous women living in Guatemala. Our data reinforce the need to strengthen the rural health system in Guatemala to deliver high-quality care not only for maternal and child health conditions but also non-communicable diseases such as diabetes.

**Table 1.**
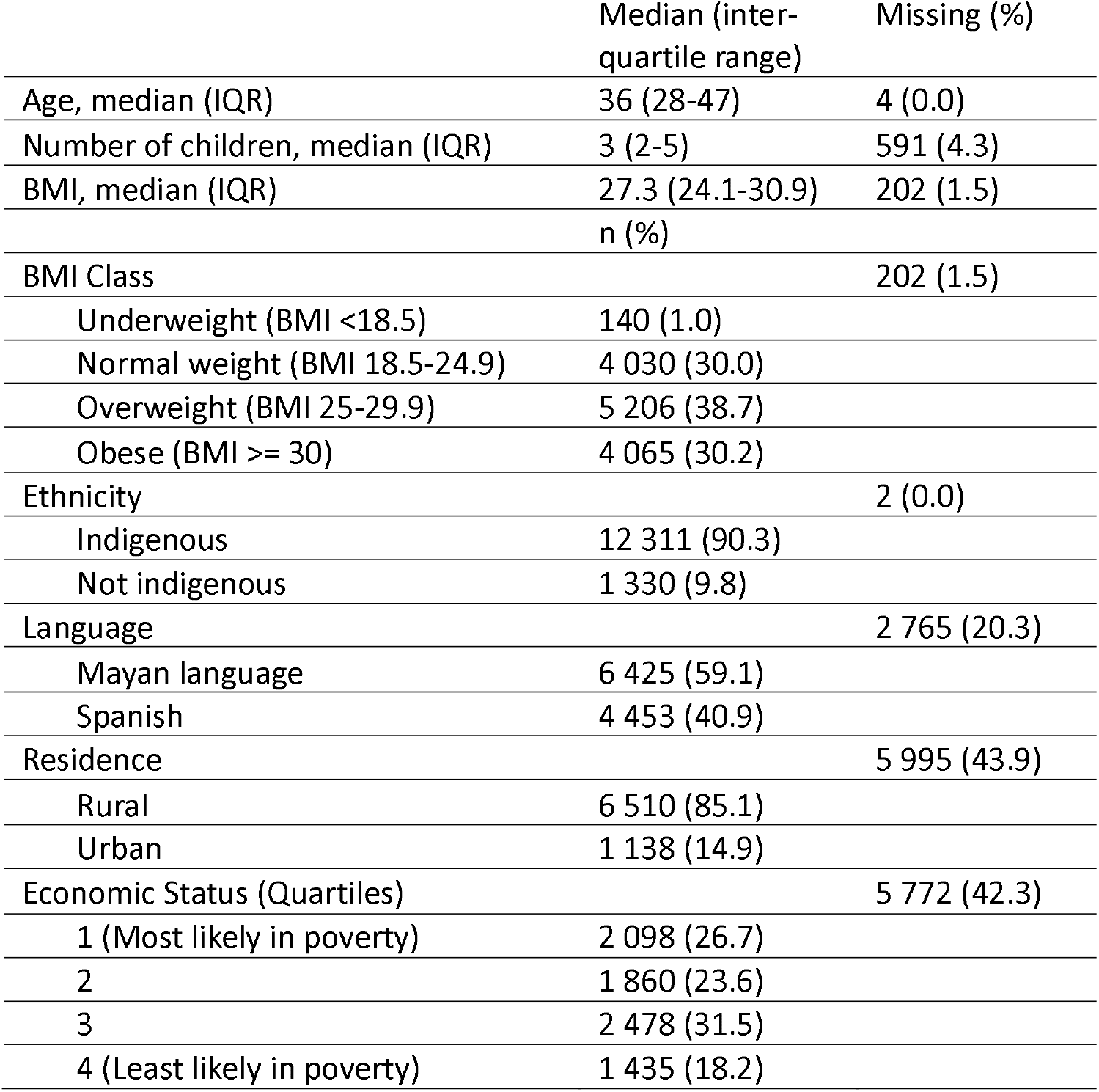
Demographic characteristics of a cohort of 13 643 primarily Indigenous women in Guatemala.

|  | Median (inter-quartile range) | Missing (%) |
| --- | --- | --- |
| Age, median (IQR) | 36 (28-47) | 4 (0.0) |
| Number of children, median (IQR) | 3 (2-5) | 591 (4.3) |
| BMI, median (IQR) | 27.3 (24.1-30.9) | 202 (1.5) |
|  | n (%) |  |
| BMI Class |  | 202 (1.5) |
| Underweight (BMI <18.5) | 140 (1.0) |  |
| Normal weight (BMI 18.5-24.9) | 4 030 (30.0) |  |
| Overweight (BMI 25-29.9) | 5 206 (38.7) |  |
| Obese (BMI ≥ 30) | 4 065 (30.2) |  |
| Ethnicity |  | 2 (0.0) |
| Indigenous | 12 311 (90.3) |  |
| Not indigenous | 1 330 (9.8) |  |
| Language |  | 2 765 (20.3) |
| Mayan language | 6 425 (59.1) |  |
| Spanish | 4 453 (40.9) |  |
| Residence |  | 5 995 (43.9) |
| Rural | 6 510 (85.1) |  |
| Urban | 1 138 (14.9) |  |
| Economic Status (Quartiles) |  | 5 772 (42.3) |
| 1 (Most likely in poverty) | 2 098 (26.7) |  |
| 2 | 1 860 (23.6) |  |
| 3 | 2 478 (31.5) |  |
| 4 (Least likely in poverty) | 1 435 (18.2) |  |

**Figure 1.**
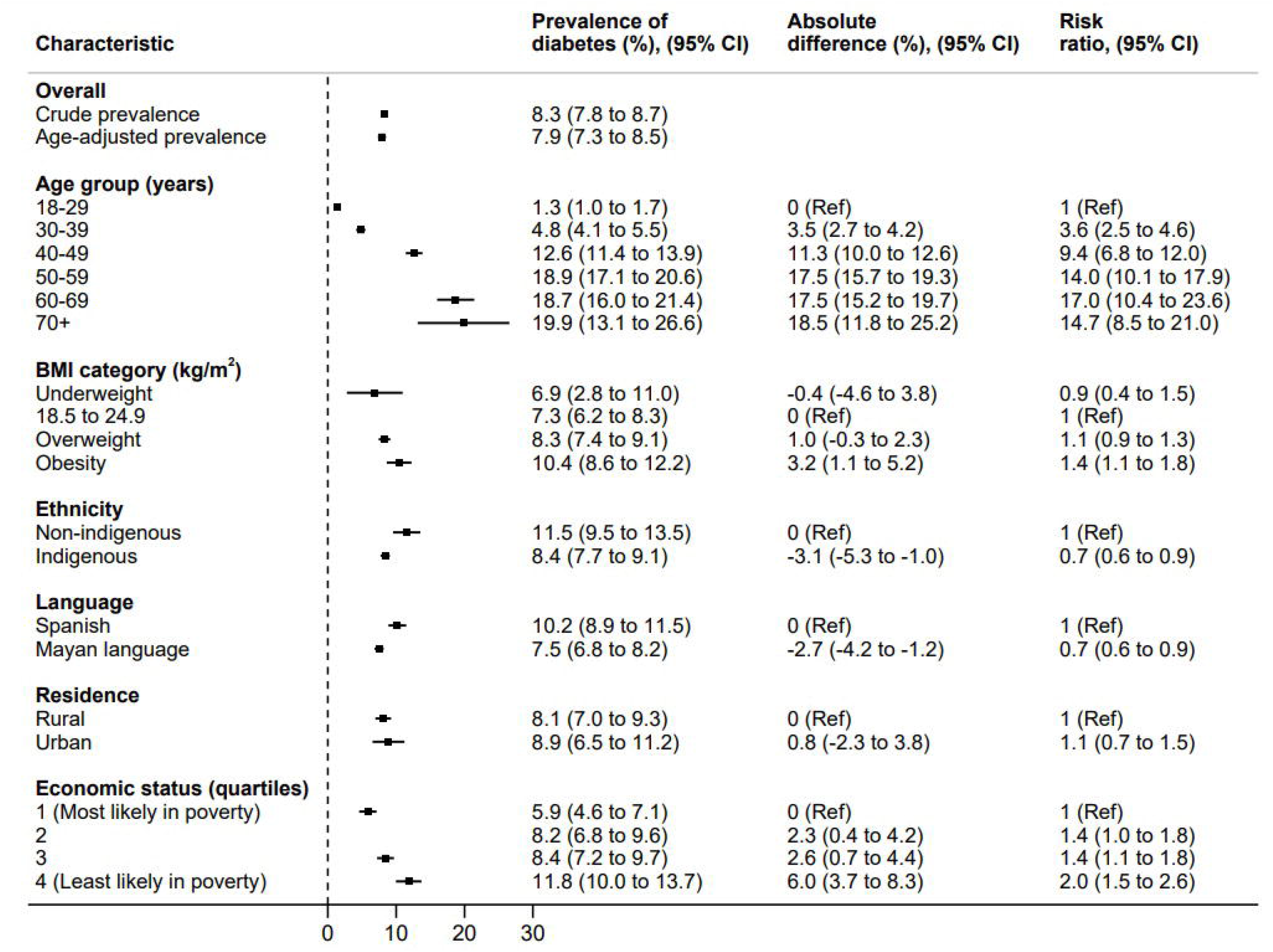
Prevalence of diabetes among a cohort of 13 643 primarily Indigenous women in Guatemala, overall and by demographic characteristic [Figure not embedded, see separate file entitled “Figure 1.jpg”] For economic status quartiles, the probabilities of poverty were as follows: quartile 1, ≥78.2%; quartile 2, ≥46.6% to <78.1%; quartile 3, ≥15.6% to <46.5%; quartile 4, <15.6%.

## Author contributions

PR and DF conceptualized the study with input from SA. PR extracted data from relevant sources. SA and PR cleaned the data and sorted variables. SA ran statistical analyses with oversight from DF and PR. SA and DF created tables and figures and drafted the manuscript. JB, YJM and CS collected the data, reviewed the work and provided feedback. All authors reviewed the results and approved the final article.

## Data availability statement

The datasets generated during and/or analyzed during the current study are not publicly available as they are proprietary data of the collaborating microfinance institution but may be made available from the corresponding author with permission from the institution upon reasonable request. Statistical code is available at the Harvard Dataverse (https://doi.org/10.7910/DVN/GA8JEA).

